# Mortality Risk and Treatment Disparities in the Chinese SMuRF-less STEMI Patients: A Nationwide Cohort Study

**DOI:** 10.1101/2025.04.08.25325498

**Authors:** Bin Sun, Chunpo Liang, Pengfei Sun, A Geru, Linjie Li, Liang Zhao, Piao Li, Yongle Li, Roger Sik-Yin Foo, Mark Yan-Yee Chan, Qing Yang, Xin Zhou

**Affiliations:** Department of Cardiology, Tianjin Medical University General Hospital, Tianjin Medical University, Tianjin 300052, China; Department of Cardiology, National University Heart Centre, 5 Lower Kent Ridge Rd, Singapore 119074, Singapore; Cardiovascular Research Institute, National University Health System, 14 Medical Drive, Singapore 117599, Singapore; Yong Loo-Lin School of Medicine, National University of Singapore, 1E, Kent, Ridge Road, Singapore 119228, Singapore

**Keywords:** ST-segment elevation myocardial infarction, Standard modifiable cardiovascular risk factors, Chinese population, Cardiovascular events, Sex differences

## Abstract

**Aims:** In ST-segment elevation myocardial infarction (STEMI) patients, standard modifiable cardiovascular risk factors (SMuRFs) are strongly associated with increased risk of morbidity and mortality. However, clinical studies have shown that SMuRF-less patients have worse clinical outcomes. Limited research exists in China, and the clinical characteristics and prognosis of SMuRF-less patients remain unclear. This study aims to provide insight into the clinical features, treatments, and outcomes of SMuRF-less patients in China.

**Methods:** We retrospectively analysed adult patients with STEMI using data from the China Chest Pain Center database. We examined the impact of SMuRF-less on short-term (0 to 30 days) and long-term (31 to 365 days) mortality. Cox-proportional hazard models and Kaplan-Meier analysis for short-term and long-term mortality.

**Results:** Of the 379,811 patients, 87,830 (23.1%) were SMuRF-less. Patients in the SMuRF-less group were older (65 years vs. 63 years), a higher proportion of female (29.4% vs. 23.3%) and less like to receiving reperfusion therapy (81.1% vs. 85.6%). On short-term SMuRFs patients had significantly lower all-cause mortality (5.1% vs 8.7%, hazard ratios (HR) 0.82 [95% Confidence interval (CI) 0.80–0.84], *P* < 0.001). However, on long-term there was no significant difference in mortality between the groups (3.4% vs 3.8%, HR: 1.00, 95% CI: 0.96-1.05; *P* = 0.841).

**Conclusions:** SMuRF-less patients exhibited higher mortality, particularly within the first 30 days after STEMI onset, with a more pronounced effect observed in male patients. Treatment disparities and delayed care were key contributors to this difference.

**Graphical abstract:** 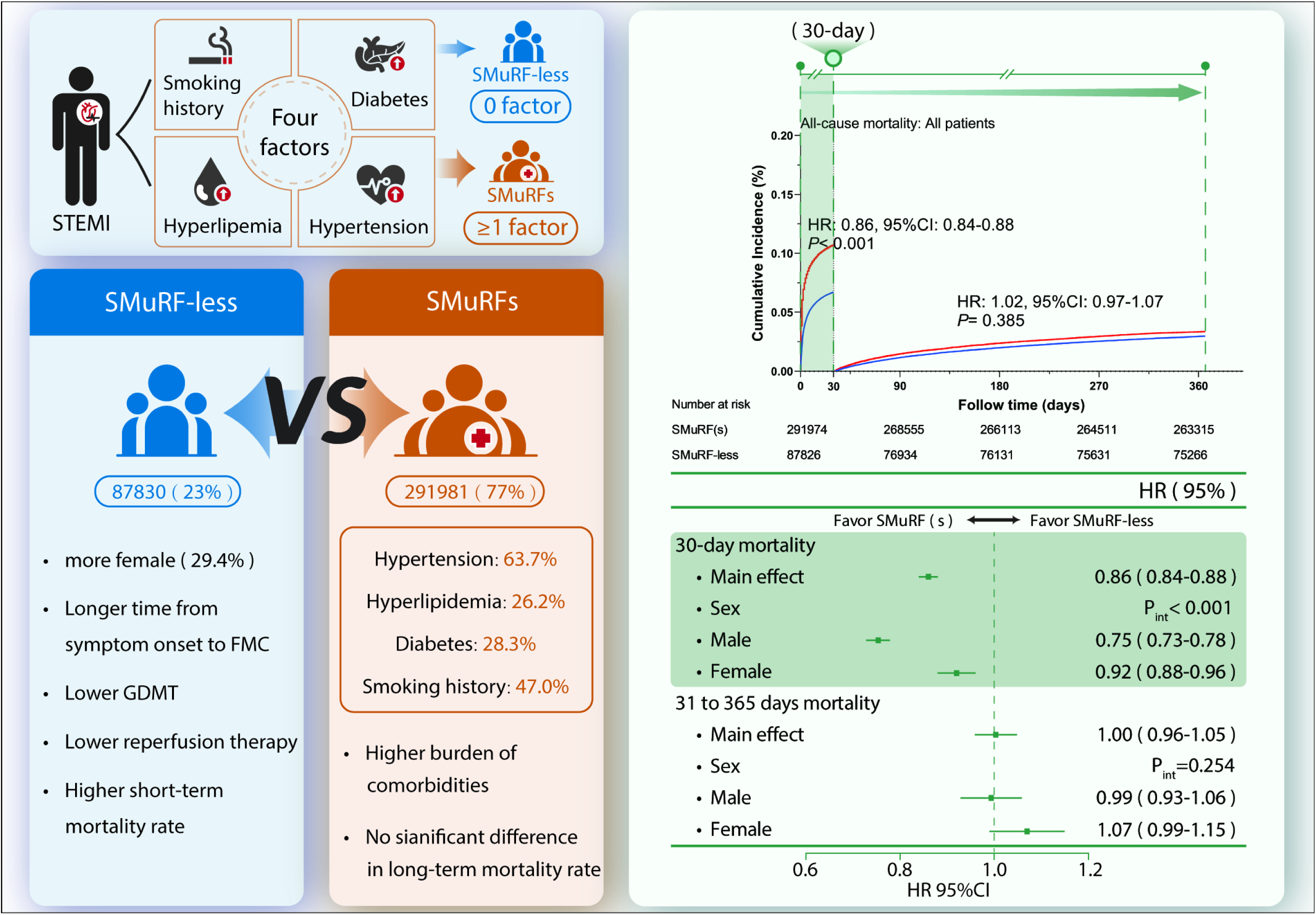

Comparison of clinical characteristics and outcomes between STEMI patients with SMuRF-less and SMuRFs. SMuRF-less was defined as having no traditional risk factors (hypertension, hyperlipidemia, diabetes, or smoking history), while SMuRFs had ≥1 risk factor. Among STEMI patients, SMuRF-less patients, who accounted for 23% of the cohort, had a higher proportion of females, longer delays to first medical contact (FMC), lower rates of guideline-directed medical therapy (GDMT) and reperfusion, and higher short-term mortality compared to SMuRFs (77%). Kaplan-Meier analysis shows lower 30-day mortality in SMuRF-less patients (HR:0.82, 95% CI: 0.80–0.84, *P* < 0.001), with no significant difference in long-term mortality (HR: 1.00, 95% CI: 0.96–1.05, *P* = 0.841).

Abbreviations: STEMI, ST elevation myocardial infarction; SMuRFs, standard modifiable cardiovascular risk factors; FMC, first medical contact; GDMT, guideline-directed medical therapy; HR, hazard ratio; CI, confidence interval.

## Introduction

The past 30 years have witnessed a rapid increase in cardiovascular disease (CVD) incidence, posing significant challenges to global healthcare systems.^1^ The 2022 Report on Cardiovascular Health and Diseases in China highlights the ongoing rise in CVD prevalence and mortality, estimating 330 million individuals currently affected and attributing two out of five deaths to CVD.^2^ Hypertension, hyperlipidemia, diabetes, obesity, and smoking are well-established modifiable cardiovascular risk factors (SMuRFs) commonly linked to CVD and routinely used to assess AMI risk in patients.^3^ Harmonized individual-level data from a global cohort showed that 22.2% and 19.1% of deaths from any cause among women and men, respectively, may be attributable to five modifiable risk factors.^4^

However, a substantial proportion of STEMI patients present without these traditional risk factors, known as SMuRF-less patients.^5^ In Western populations, studies have shown that SMuRF-less patients tend to have higher early mortality and worse short-term outcomes, despite being perceived as low-risk due to the absence of modifiable risk factors.^6^ These patients often experience delays in diagnosis, treatment initiation, and lower adherence to guideline-directed therapies, which contribute to their adverse outcomes.^6, 7^ Similar findings have been observed in several countries, including Canadian and US cohorts, where SMuRF-less patients exhibit higher in-hospital mortality and poorer prognoses.^8, 9^

Studies from developing countries are scarce, with China having only one study based on data from 2014.^10^ However, research on this population remains limited, and there is a lack of large-scale, nationwide studies examining their clinical characteristics, treatment disparities, and outcomes. Previous studies have identified underutilization of evidence-based therapies, particularly among women, contributing to the increased mortality observed in this group.^6, 11^ Despite this, most research in China has been regional or hospital-based, with few exploring long-term outcomes or addressing the impact of sex and treatment delays on survival.

This present study aims to provide a comprehensive analysis of the clinical features, treatment patterns, and short-term and long-term mortality of SMuRF-less STEMI patients in China, using data from the China Chest Pain Center database, one of the largest national registries in the country. By comparing SMuRF-less and SMuRFs patients, we sought to highlight the critical need for timely interventions and improved adherence to guideline-directed therapies, particularly for vulnerable subgroups like women, to improve overall clinical outcomes.

## Methods

### Study design and population

The National Chest Pain Centers Program is a large-scale, multifaceted initiative focused on continuous quality improvement for managing acute chest pain. In October 2019, the National Health Commission launched the China Alliance of Chest Pain Centers (CPC) to enhance accreditation standards and expand the establishment of chest pain centers nationwide. The program operates through a centralized, web-based registry, the Chinese Cardiovascular Association (CCA) Database-Chest Pain Center, where hospitals independently collect and submit case data. These data, extracted from medical records, cover patient demographics, pre-hospital interventions, presenting symptoms, in-hospital treatments (including medication and reperfusion), and discharge outcomes.^12^ By the end of 2021, 5,107 hospitals participated in the accreditation project, with 2,096 achieving accreditation—1,047 under the standard model and 1,049 under the basic model.

We analyzed all patient records from accredited Chest Pain Centers stored in the database, covering admissions from January 1, 2017, to December 31, 2021. Eligible patients for this analysis were adults aged 18 years or older who presented with suspected acute coronary syndrome and were diagnosed with STEMI. Exclusion criteria included a history of revascularization procedures (such as percutaneous coronary intervention [PCI], coronary artery bypass grafting [CABG], or myocardial infarction) and unclear SMuRFs group assignments (**Figure 1**).

**Figure 1.**
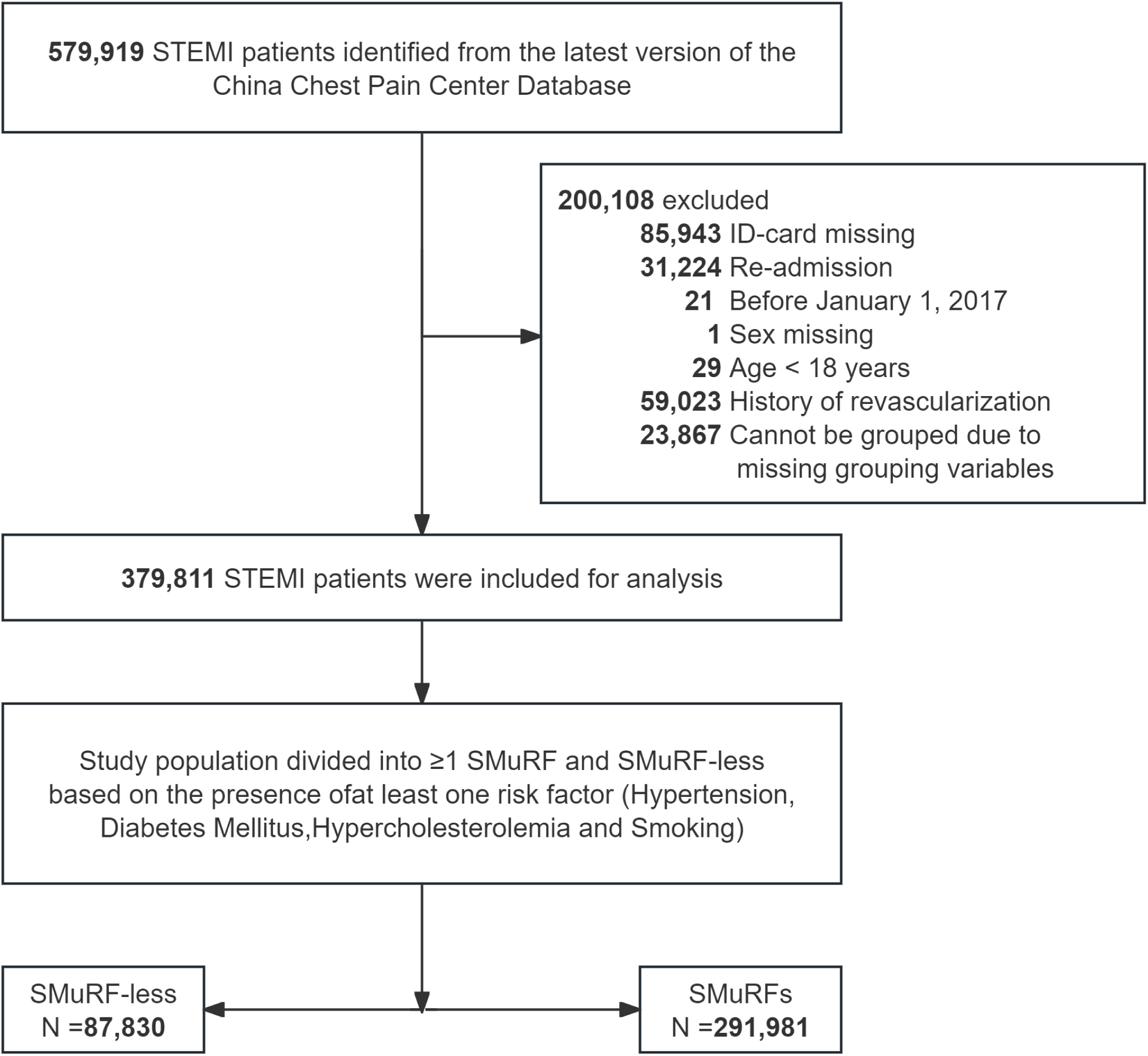
Flow-chart of patients included in the study. Abbreviations: ID, identity document; STEMI, ST elevation myocardial infarction; SMuRFs, standard modifiable cardiovascular risk factors.

The data were processed to remove all information that could reveal patients’ identity. Anonymized data are available through a formal application process and after review by the Data Management Committee of the CCA Database-Chest Pain Center. The study protocol was conducted in accordance with the Declaration of Helsinki (as revised in 2013) and approved by the institutional review board at the Tianjin Medical University General Hospital (IRB2022-WZ-145). Informed consent was waived due to the retrospective nature of the study.

### Definition of SMuRFs

The exposure variable was defined as the presence of one or more SMuRFs: a history of hypertension, diabetes, hyperlipidemia, or smoking. Hypertension, diabetes, and hyperlipidemia histories were classified based on diagnoses from the hospital. A history of smoking was defined as regular smoking (≥1 cigarette/day) before the index hospitalization. SMuRFs patients were defined as having at least one of the aforementioned risk factors, while SMuRF-less patients had no documented SMuRFs in their admission medical records.

### Study variables

Reperfusion therapy was defined as the reopening of the occluded coronary vessel and included thrombolysis, primary PCI, transport PCI, and CABG. Indicators of STEMI patients’ access to care include: time from symptom onset to first medical contact (S-to-FMC), time from first medical contact to wire crossing (FMC-to-W) in primary PCI patients, and time from first medical contact to needle initiation (FMC-to-N) in patients receiving thrombolytic therapy. They also include metrics of effective reperfusion, such as successful thrombolysis or achieving a Thrombolysis in Myocardial Infarction (TIMI) flow grade of 3, and timely reperfusion, defined as FMC-to-N <30 minutes or FMC-to-W <120 minutes. We also included data on patients’ admission blood pressure, heart rate, Killip classification, in-hospital treatments (including reperfusion), and emergency medications administered at admission.

The primary outcome was all-cause mortality assessed at two landmark periods: short-term (0 to 30 days) and long-term (31 to 365 days), defined as death within the specified time from the index date or period start. The secondary outcome was cardiac mortality. The study utilized unique national IDs to link patient data with the China National Death Registration System,^13^ ensuring accurate vital status tracking for all patients in the CPC database.

### Statistical analysis

Categorical variables were summarized as frequencies and percentages. Numerical variables were summarized as the mean (standard deviation, SD) or the median (interquartile range, IQR), depending on data distribution. To explore differences in characteristics, in-hospital findings, and management between SMuRF-less patients and patients with SMuRFs, categorical variables were analyzed using the χ² test, and continuous variables using Student’s t-tests. Associations between the exposure variable and study outcomes were assessed using multivariable Cox egression models to calculate hazard ratios (HR) and 95% confidence intervals (CI). Cumulative mortality risk, stratified by SMuRF-less status, was constructed using the Kaplan-Meier method. The proportional hazards assumption was tested using the Schoenfeld residuals test. Due to violation of the proportional hazard assumption, Kaplan-Meier curves were compiled in the context of a landmark analysis with a 30 days threshold to assess at short-term (0 to 30 days) and long-term (31 to 365 days) periods, defined as death within the specified time from the index date or period start. Covariates for adjustment included: sex, age, pulse, respiration rate, blood pressure on admission, previous medical history, Killip class, reperfusion therapy, and initial in-hospital medications. We applied multiple imputations using chained equations across five datasets to impute missing values for these clinical variables.

We evaluated the effects of SMuRFs status on short-term and long-term mortality, considering sex-specific treatment differences in STEMI patients. Additionally, we analyzed the characteristics and outcomes of SMuRFs versus SMuRF-less patients in both males and females.

All analyses were done with STATA software (version 17.0) and R software (version 4.2.0). A two-sided p value of less than 0.05 was considered to indicate statistical significance.

## Results

From January 1, 2017, to December 31, 2021, 579,901 adult STEMI patients were identified in the the China Chest Pain Center (CCPC) database. After excluding individuals with a history of myocardial infarction or revascularization, the remaining 379,811 patients constituted the study population. Of the 379,811 patients, 87,830 (23.1%) were SMuRF-less, and 291,981 (76.9%) had one or more SMuRFs before hospitalization (**Figure 1**).

Baseline demographics, distribution of SMuRFs, and in-hospital treatments are presented in **Table 1**. The median age of the study population was 64 years. Compared to SMuRF-less patients, those with SMuRFs were younger at the onset of STEMI. Of these patients, 24.4% were women. Patients with SMuRFs had a lower proportion of females. Among patients with SMuRFs, the most common condition was hypertension (185,982 [63.7%]), followed by smoking (137,210 [47.0%]), diabetes (79,927 [28.3%]), and hyperlipidemia (76,365 [26.2%]). SMuRF-less patients were less likely to have histories of conditions such as obesity, coronary heart disease (CHD), family history of premature CHD, heart failure, peripheral artery disease, and chronic kidney disease.

**Table 1.** Patient characteristics.

| Characteristics | Total<br>n=379811 | SMuRF-less<br>n=87830 | SMuRF(s)<br>n=291981 | P-value |
| --- | --- | --- | --- | --- |
| **Admission characteristics** |  |  |  |  |
| Age, years, median (IQR) | 64.0 (53.0, 72.0) | 65.0 (54.0, 74.0) | 63.0 (53.0, 72.0) | < 0.001 |
| Female, n (%) | 93949 (24.7) | 25815 (29.4) | 68134 (23.3) | < 0.001 |
| **SMuRF, n (%)** |  |  |  |  |
| Hypertension | 185982 (49.0) | 0 (0.0) | 185982 (63.7) | < 0.001 |
| Hyperlipidemia | 76365 (20.1) | 0 (0.0) | 76365 (26.2) | < 0.001 |
| Diabetes | 79927 (21.6) | 0 (0.0) | 79927 (28.3) | < 0.001 |
| Smoking history | 137210 (36.1) | 0 (0.0) | 137210 (47.0) | < 0.001 |
| **Clinical characteristics** |  |  |  |  |
| SBP, mean (SD) | 132.2 (26.9) | 126.9 (25.5) | 133.9 (27.1) | < 0.001 |
| Awareness, sober (%) | 374610 (98.7) | 86313 (98.3) | 288297 (98.8) | < 0.001 |
| Respiratory rate, median (IQR) | 20 (18, 20) | 20 (18, 20) | 20 (18, 20) | < 0.001 |
| HR, median (IQR) | 77 (65, 90) | 76 (64, 89) | 77 (65, 90) | < 0.001 |
| Killips I class, n (%) | 270705 (74.7) | 61803 (73.5) | 208902 (75.1) | < 0.001 |
| Killips IV class, n (%) | 21823 (6.0) | 5849 (7.0) | 15974 (5.7) | < 0.001 |
| **Previous history, n (%)** |  |  |  |  |
| Family history of premature CHD | 9908 (2.6) | 1676 (1.9) | 8232 (2.8) | < 0.001 |
| Obesity | 24219 (6.4) | 1860 (2.1) | 22359 (7.7) | < 0.001 |
| Coronary heart disease | 122419 (32.2) | 23263 (26.5) | 99156 (34.0) | < 0.001 |
| Atrial fibrillation | 14473 (3.8) | 3098 (3.5) | 11375 (3.9) | < 0.001 |
| Heart failure | 20652 (6.0) | 4076 (5.0) | 16576 (6.3) | < 0.001 |
| Valvular heart disease | 4623 (1.2) | 982 (1.1) | 3641 (1.2) | 0.002 |
| Peripheral arterial disease | 12813 (3.5) | 1756 (2.0) | 11057 (3.9) | < 0.001 |

Table 1. Continued
| Characteristics | No. of Patients<br>n=379811 | SMuRF-less<br>n=87830 | SMuRF(s)<br>n=291981 | P-value |
| --- | --- | --- | --- | --- |
| COPD | 7362 (2.0) | 1413 (1.6) | 5949 (1.0) | < 0.001 |
| CKD | 11759 (3.2) | 1389 (1.6) | 10370 (3.7) | < 0.001 |
| Anemia | 9398 (2.5) | 2056 (2.3) | 7342 (2.6) | < 0.001 |
| Peptic ulcer | 6688 (1.8) | 1238 (1.4) | 5450 (1.9) | < 0.001 |
| Thyroid dysfunction | 5810 (1.6) | 945 (1.1) | 4865 (1.7) | < 0.001 |
| Cancer | 3945 (1.1) | 951 (1.1) | 2994 (1.1) | 0.54 |
| **Emergency medication, n (%)** |  |  |  |  |
| Antiplatelet | 338650 (93.4) | 77025 (91.5) | 261625 (94.0) | < 0.001 |
| Statin | 273822 (79.6) | 61495 (76.9) | 212327 (80.4) | < 0.001 |
| β-blocker | 119819 (35.7) | 25327 (32.4) | 94492 (36.7) | < 0.001 |
| **In-hospital management, n (%)** |  |  |  |  |
| S-to-FMC | 372882 | 86059 | 286823 | NA |
| Median (IQR), min | 167 (69, 547) | 173 (70, 578) | 165 (68, 540) | < 0.001 |
| FMC-to-W | 184194 | 36580 | 147614 | NA |
| Median (IQR), min | 93 (70, 151) | 93 (70, 153) | 93 (70, 151) | 0.55 |
| FMC-to-N | 46878 | 11995 | 34883 | NA |
| Median (IQR), min | 34 (25, 62) | 33 (24, 60) | 35 (25, 63) | < 0.001 |
| Effective reperfusion | 184342 (83.9) | 37354 (79.0) | 146988 (85.3) | < 0.001 |
| Timely reperfusion | 141272 (64.2) | 29321 (63.1) | 111951 (64.4) | < 0.001 |
| Reperfusion therapy | 306862 (84.6) | 68263 (81.1) | 238599 (85.6) | < 0.001 |
| Primary PCI | 219391 (60.6) | 44993 (53.5) | 174398 (62.7) | < 0.001 |
| Thrombolysis | 38351 (10.6) | 10324 (12.3) | 28027 (10.1) | < 0.001 |

Abbreviations: SMuRFs, standard modifiable cardiovascular risk factors; SBP, systolic blood pressure; HR, heart rate; CHD, coronary heart disease; COPD, chronic obstructive pulmonary disease; CKD, chronic kidney disease; PCI, percutaneous coronary intervention; S-to-FMC, symptom onset-to-first medical contact; FMC-to-W,
first medical contact-to-wise; FMC-to-N, first medical contact-to-needle.

Upon presentation of STEMI, SMuRF-less patients had significantly lower systolic blood pressure and heart rate compared to those with SMuRFs, and the cardiac function classification at admission was worse. The S-to-FMC and FMC-to-N times were significantly shorter in patients with SMuRFs. However, there was no significant difference in FMC-to-W between SMuRF-less patients and patients with SMuRFs. Patients with SMuRFs were more likely to achieve effective reperfusion (85.3% vs. 79.0%).

In terms of emergency medication therapy, SMuRF-less patients were less likely to receive Guideline-Directed medical therapy, included antiplatelet drugs (91.5% vs. 94.0%), statins (76.9% vs. 80.4%), and β-blockers (32.4% vs. 36.7%). Furthermore, SMuRF-less patients were less likely to undergo reperfusion therapy (81.1% vs. 85.6%) and primary PCI (53.5% vs. 62.7%).

### Short-term and Long-term mortality

During the 1-year follow-up, a total of 40,689 patients died, including 12,365 (14.1%) in the SMuRF-less group and 28,324 (9.7%) in the SMuRFs group. The curves diverged from the initial day of STEMI presentation. The cumulative mortality risk is depicted in **Figure 2A**.

**Figure 2.**
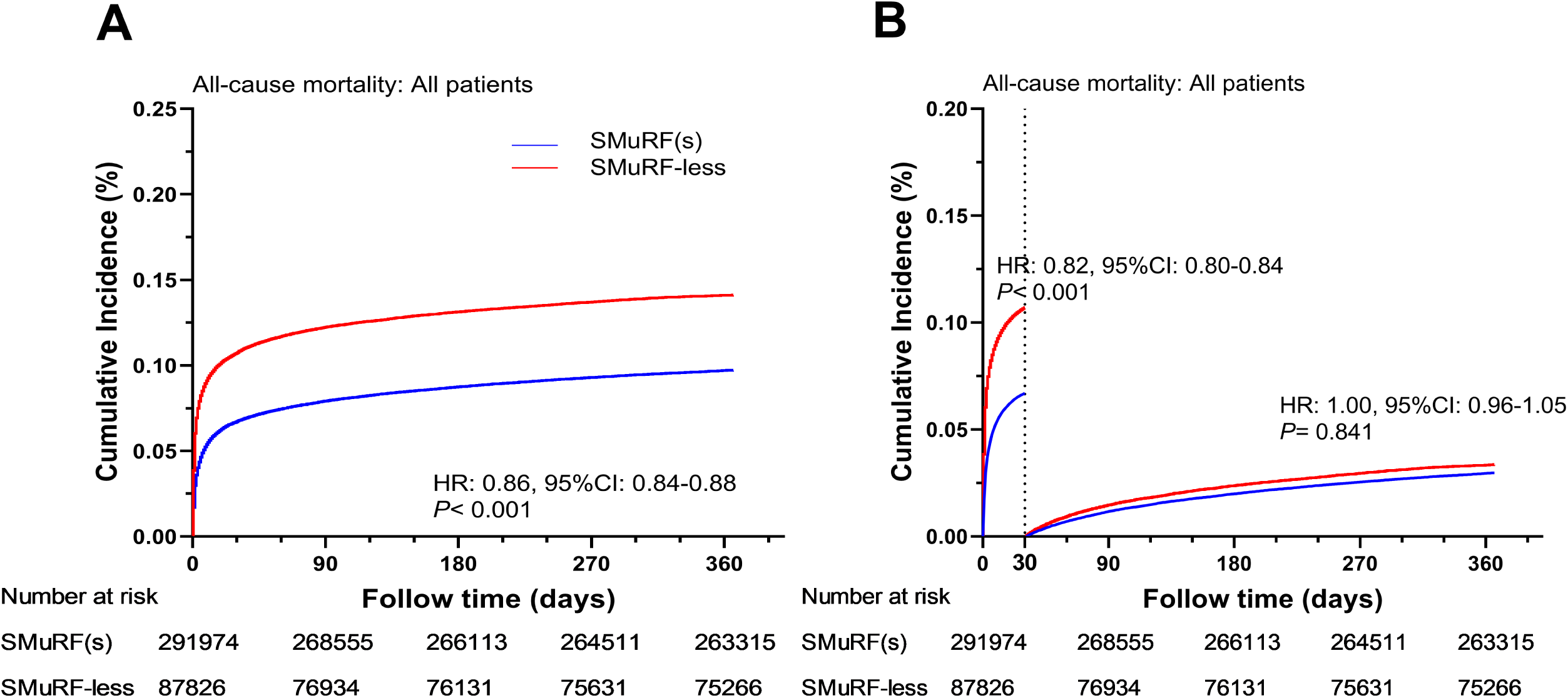
Kaplan-Meier survival curves for all-cause mortality up to 1 year. A): The Kaplan-Meier curve for all-cause mortality within 1 year in the overall population. B): The landmark analysis curve for all-cause mortality within 1 year in the overall population. Abbreviations: SMuRFs, standard modifiable cardiovascular risk factors.

For 30-day mortality (**Figure 2B and Figure 3**), 9,377 out of 87,626 patients (10.7%) in the SMuRF-less group died, compared to 19,546 out of 291,571 patients (6.7%) in the SMuRFs group. SMuRFs status was associated with a lower risk of death in the crude analysis (HR: 0.61, 95% CI: 0.60-0.63; *P* < 0.001) and in the multivariable-adjusted analysis (HR: 0.82, 95% CI: 0.80-0.84; *P* < 0.001).

**Figure 3.**
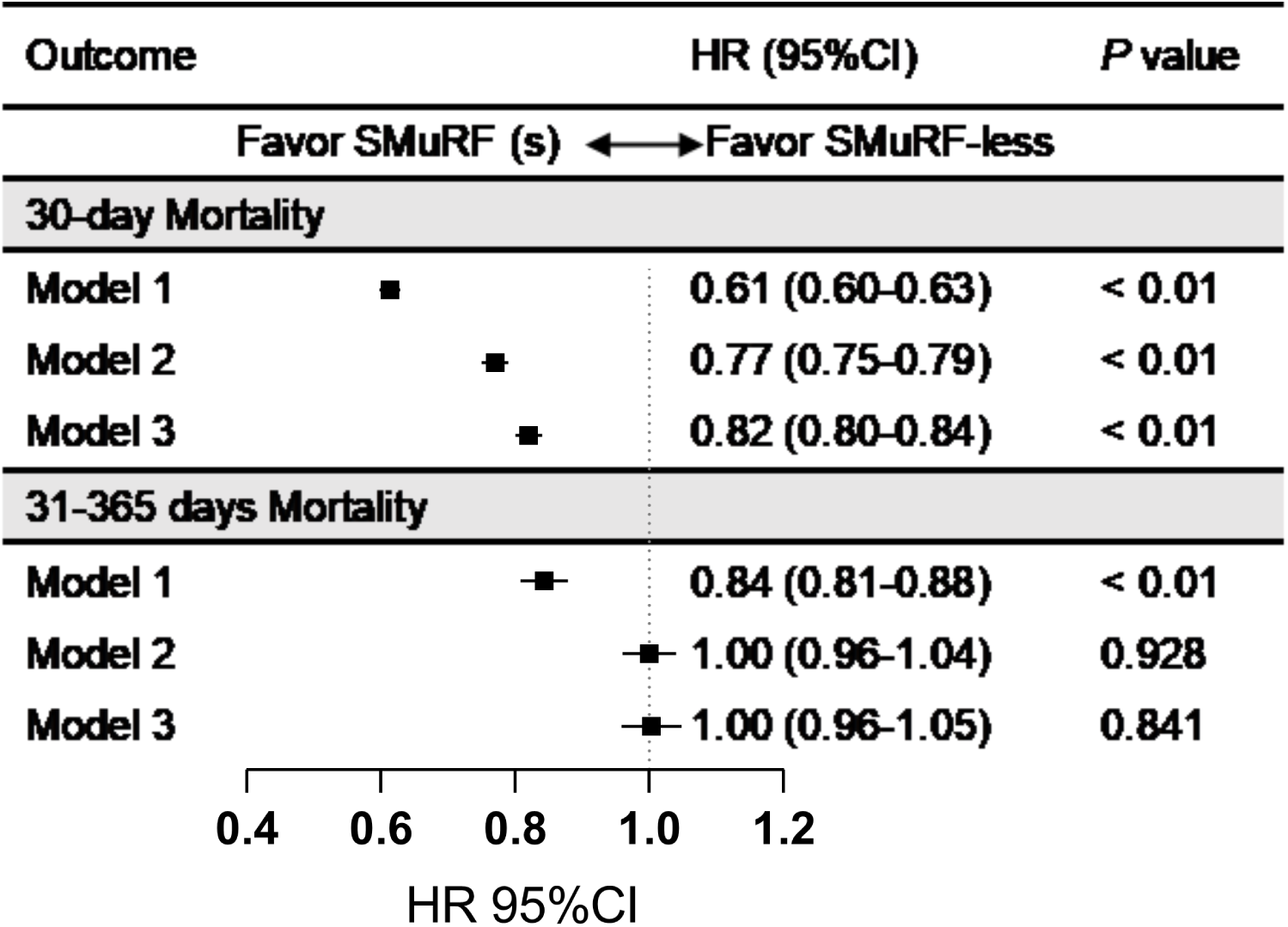
HRs for all-cause mortality between SMuRF-less patients and patients with SMuRFs within 30 days and 31 to 365 days. Point estimates and 95% CIs are presented from Cox regression analyses in the total population. Cox regression models, Model 1: Unadjusted; Model2: Model adjusted for admission sign (age, sex, respiratory rate, heart rate, awareness, and Killips classific ation on admission); Model 3: Model adjusted for admission sign, past history (CHD, family history of premature CHD, atrial fibrillation, heart failure, valvular heart disease, peripheral arterial disease, COPD, CKD, anemia peptic ulcer, thyroid dysfunction, cancer) and treatment measures (statins, betablocker, antiplatelet, reperfusion therapy, symptom onset-to-first medical contact). Abbreviations: SMuRFs, standard modifiable cardiovascular risk factors; HR, hazard ratio; CHD, Coronary heart disease; COPD, chronic obstructive pulmonary disease; CKD, chronic kidney disease.

For mortality from 31 to 365 days (**Figure 2B and Figure 3**), 2,988 out of 78,249 patients (3.8%) in the SMuRF-less group died, compared to 8,778 out of 272,025 patients (3.2%) in the SMuRFs group. In crude analysis without covariate adjustment, patents with SMuRFs had a lower mortality risk (HR: 0.84, 95% CI: 0.81-0.88; *P* < 0.001). However, in the multivariable-adjusted analysis, SMuRFs status was not associated with increased mortality (HR: 1.00, 95% CI: 0.96-1.05; *P* = 0.841).

In the secondary outcome analysis, within 30 days of admission, cardiac death accounted for 80.4% (23,209/28,923) of all causes of death, and at 1-year follow-up, cardiac death accounted for 78.3% (31,846/40,689) of all causes of death. The results for cardiac death were similar to those for all-cause mortality. (**Figure S1 and Figure S2**)

### Subgroup analysis

However, interactions were observed only for age and sex with respect to 30-day mortality (**Figure 4**). During the 30-day follow-up period, we observed that while mortality increased with age, the effect of SMuRF-less status on mortality gradually diminished (*P* for interaction <0.001, **Figure 4**). By the 31 to 365 days follow-up, the mortality difference between the SMuRFs and SMuRF-less groups disappeared. Among patients with a family history of premature CHD, the impact of SMuRF-less status on mortality was no longer observed, either within 30 days or between 31 to 365 days (**Figure 4**).

**Figure 4.**
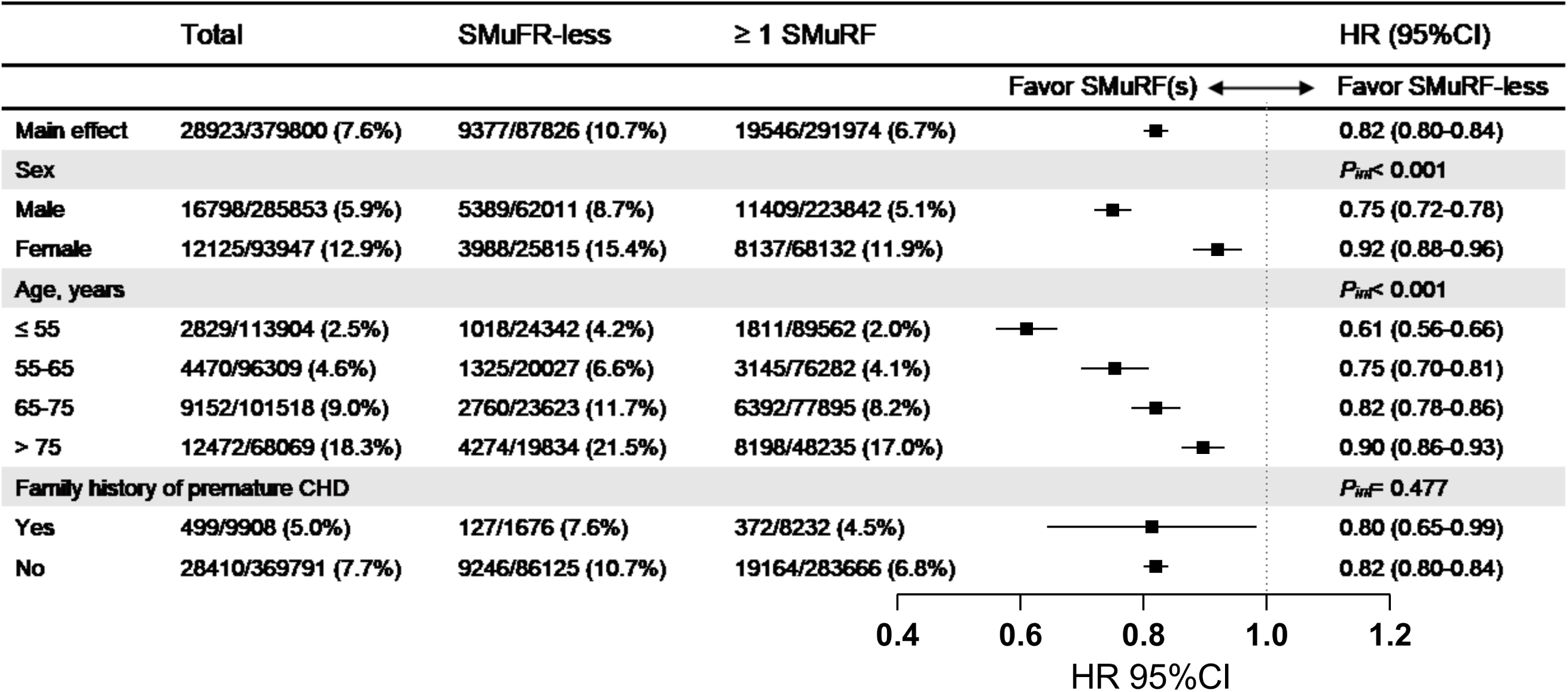
Subgroup analysis of HRs for primary outcome between SMuRF-less patients and patients with SMuRFs with 30 days. The subgroup analysis of point estimates and 95% confidence intervals (CIs) derived from Cox regression analyses for all-cause mortality within 30 days in the total population. Cox regression analyses, Model: Model adjusted for admission sign, past history and treatment measures. Abbreviations: SMuRFs, standard modifiable cardiovascular risk factors; CHD, coronary heart disease; HR, hazard ratio.

Notably, among male SMuRFs patients, after adjusting for covariates, those with SMuRFs had significantly lower mortality risk (HR: 0.75, 95% CI: 0.72–0.78; *P* < 0.001). The outcomes in women were similar, though the effect was weaker: after adjustment, significant difference in mortality was observed for patients with SMuRFs (HR: 0.92, 95% CI: 0.88–0.96; *P* < 0.001). The impact of SMuRF-less status on 30-day mortality disappeared in women (*P* for interaction < 0.001; **Figure 4**). Surprisingly, the effect of SMuRF-less status on 1-year mortality was no longer evident in either male or female STEMI patients after excluding those who died within 30 days (**Figures S3**). Baseline demographics, SMuRFs distribution, and in-hospital treatments, stratified by sex and SMuRFs status, are shown in Table S1. Crude and adjusted all-cause and cardiac mortality differences between the SMuRF-less and SMuRFs groups are presented in **Figure 5 and Figure S1**.

**Figure 5.**
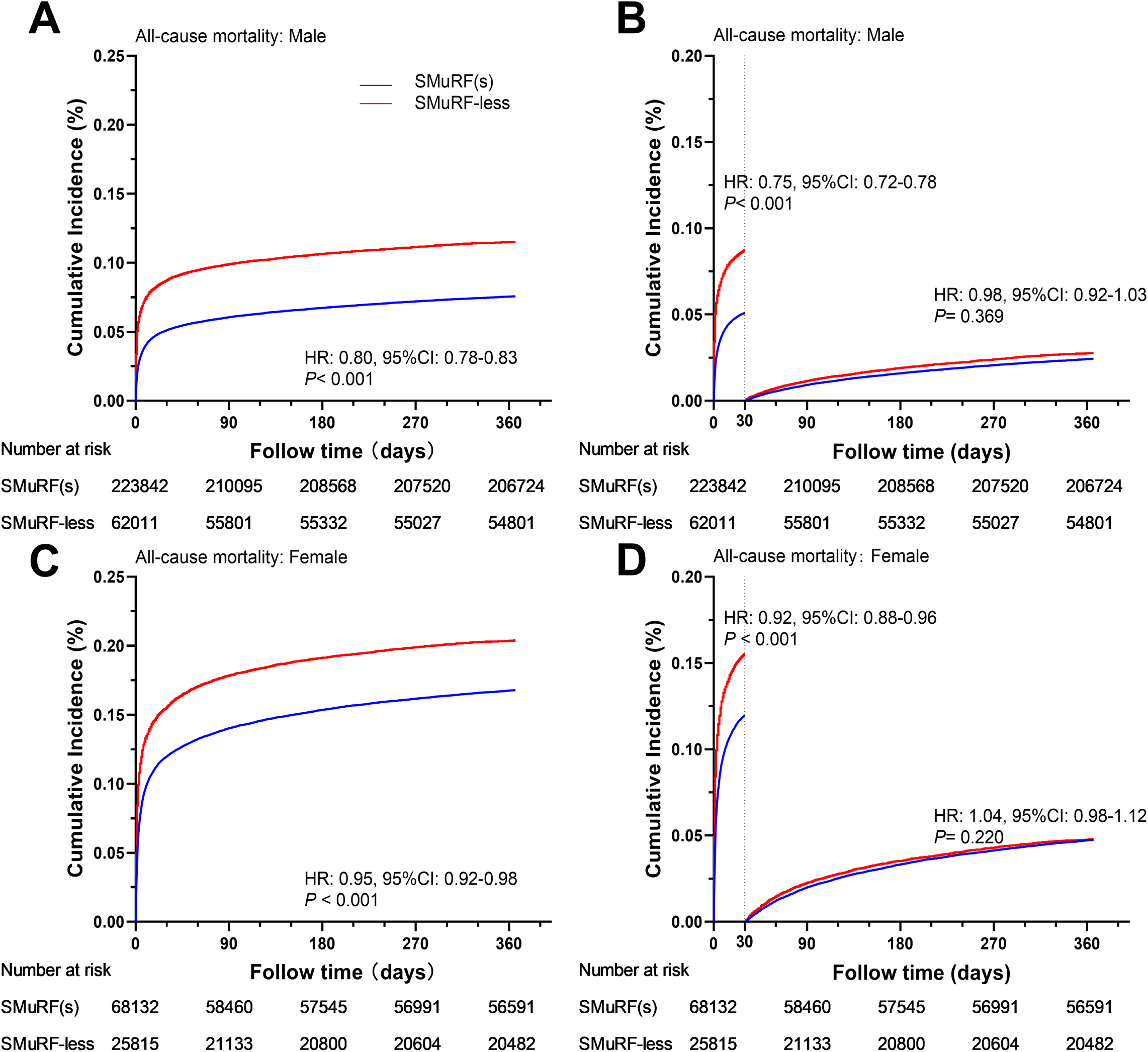
Sex subgroup analysis of Kaplan-Meier survival curves for 1-year all-cause mortality. A): The Kaplan-Meier curve for all-cause mortality within 1 year in the male patient population. B): The landmark analysis curve for all-cause mortality within 1 year in the male patient population. C): The Kaplan-Meier curve for all-cause mortality within 1 year in the female patient population. D): The landmark analysis curve for all-cause mortality within 1 year in the female patient population. Abbreviations: SMuRFs, standard modifiable cardiovascular risk factors.

## Discussion

By utilizing the CCPC dataset from 2017 to 2021, which includes over 300,000 STEMI patients from more than 2,000 centers across China, this nationwide, multicenter retrospective cohort study revealed the following findings. First, SMuRF-less patients experienced delays in treatment and were less likely to receive reperfusion therapy compared to those with SMuRFs. Second, while SMuRF-less status was associated with increased 30-day mortality, this effect diminished over the long term (31 to 365 days). Third, the detrimental impact of SMuRF-less status appeared more pronounced in males and showed age-dependent attenuation. These findings highlight the complexity of SMuRF-less patients, particularly in relation to sex-specific treatment disparities and the evolving impact of SMuRF-less status over time. (**Graphical abstract**)

In our cohort, 23.1% of patients were SMuRF-less, consistent with previous studies where the proportion of SMuRF-less patients in STEMI populations ranged from 11.0% to 26.2%.^6, 7, 14–17^ Additionally, the proportion of SMuRF-less patients has been steadily increasing in studies from certain countries.^5, 7^ Across all studies, SMuRF-less patients were older, had more comorbidities, initiated treatment later, and were less likely to receive treatment. In both the overall AMI patient population and various AMI subtypes, the proportion of females in the SMuRF-less group typically ranges from 20% to 40%. The research findings regarding the disparity in the proportion of females between the SMuRF-less and SMuRFs groups are inconsistent.^6, 7, 14^ In the SWEDHEART study in Sweden, the proportion of females in the SMuRF-less group among STEMI patients was lower than in the SMuRFs group (23.5% vs. 34.5%).^6^ Similar trends were also observed in research from the United States and Australia.^7, 17^ Interestingly, our data showed that the proportion of SMuRF-less patients was higher in females, which is consistent with previous studies from China, South Korea, Japan, and Singapore.^10, 18, 19^ This may be related to the lower smoking rates among Chinese women.^20^

Our results demonstrated a 14% lower 30-day mortality risk in the SMuRFs group compared to the SMuRF-less group in STEMI patients. Similar to most studies, AMI patients without traditional cardiovascular risk factors exhibited a significantly higher risk of mortality, a difference that remained after adjusting for baseline characteristics.^6, 9, 19^ A meta-analysis of 1.28 million ACS patients found that the risk of in-hospital mortality was 1.57 times higher in the SMuRF-less group compared to the SMuRFs group.^21^ However, some studies have reported no significant difference in mortality between the two groups, or even a lower risk of death in the SMuRF-less group. For instance, a Korean registry of 11,390 AMI patients treated with PCI found no difference in mortality between the groups.^18^

Our study’s results were consistent with those of most studies, but the significant difference in mortality between the SMuRF-less and SMuRFs groups appeared early, as indicated by the KM curve. Subsequently, a landmark analysis was performed to categorize follow-up time into two periods: within 30 days after admission and between 31 to 365 days after admission. The results aligned with our expectations: mortality in the SMuRF-less group was significantly higher than in the SMuRFs group within 30 days of admission, and this difference remained significant after adjusting for multiple risk factors. However, between 31 to 365 days after admission, the mortality difference between the groups disappeared. This further supports the notion that the higher mortality in SMuRF-less patients occurs early in the disease.

All studies showed an older age of onset in the SMuRF-less group.^6, 14, 21, 22^ Age may directly influence the baseline risk of CHD, independent of other risk factors.^23^ One possible explanation is that in patients without traditional risk factors, disease typically manifests at older ages, when the baseline risk has increased enough to precipitate the condition.^24^ SMuRF-less patients are more likely to develop cardiogenic shock, potentially due to the absence of risk factors such as hypertension and diabetes, which may result in a lack of myocardial ischemic preconditioning and reduced tolerance.^22, 25^ Some studies have also shown that the myocardial infarct size is larger in the SMuRF-less group.^26^

Studies have reported mixed results regarding delays in seeking care. Our study clearly shows that patients in the SMuRF-less group experience significant delays in seeking medical attention, possibly due to a lack of preventive awareness. The SWEDEHEART study reported a shorter time from symptom onset to hospital admission in the SMuRF-less group (3.0 hours vs 3.1 hours).^6^ However, Swiss and Japanese studies did not find any difference in delay between the two groups.^27, 28^ Regarding in-hospital delays, the “door-to-balloon” time was longer for SMuRF-less patients in the Japanese study (90 minutes vs. 82 minutes, P=0.04).^28, 29^ Encouragingly, no in-hospital delay was observed in our study, and no significant difference in FMC-to-W was found between the two groups. Consistent with previous studies, in-hospital use of aspirin, statins, or β-blockers was less frequent in the SMuRF-less group than in the SMuRFs group,^9, 14^ despite guidelines^30, 31^ recommending early administration of these drugs in the early stages of disease, unless contraindicated.

In our study, we found that SMuRF-less status was associated with increased 30-day mortality in both male and female STEMI patients. Interestingly, the 30-day mortality rate in female SMuRF-less patients was nearly twice that of male SMuRF-less patients, by contrast, the effect was weaker in female. In line with previous studies, women, especially those without traditional risk factors, should receive greater attention to guideline-recommended treatments.^6, 7, 16^

The differences in outcomes between men and women may be due to the varying impact of risk factors on each sex. Studies have shown that risk factors like hypertension, diabetes, and smoking are more strongly associated with cardiovascular disease in women.^32, 33^ In particular, smoking poses a higher risk for women, with female smokers having a 25% higher relative risk of CHDcompared to male smokers, independent of other cardiovascular risk factors. Furthermore, the pooled relative risk reduction for women compared to men increased by 2% for each additional year of follow-up.^34^ This may also explain why female patients in the SMuRF-less group had a higher risk of death than those in the SMuRFs group in Western studies.

Finally, similar to the findings in the overall population, after excluding those who died within 30 days, no significant difference in 1-year mortality was observed between SMuRF-less and SMuRFs groups, suggesting that the prognosis difference was primarily evident in the acute phase. This was consistent with findings from the SWEDEHEART study.^6^

As cardiovascular risk factor management has improved, the incidence and mortality of CHDand acute myocardial infarction have decreased. The INTERHEART study reported that cardiovascular risk factors accounted for 90% of the population attributable risk (PAR) of MI.^35^ While inflammation is recognized as a key factor in atherosclerosis, most studies show that SMuRF-less patients have lower or similar C-reactive protein levels compared to SMuRFs patients, suggesting inflammation may not be a major driver in this group.^6, 18^ In addition to traditional risk factors, genetics play a crucial role in CHD. While controlling traditional risk factors has reduced mortality, total population attributable risks remain below 100%, highlighting the need to identify new mechanisms and markers. Although polygenic risk scores (PRS) have shown limited impact compared to traditional assessments,^36^ they may support earlier CHD prevention in younger populations, particularly for those in the highest 20% of genetic risk.^37^ Cardiovascular risk assessment is recommended for identifying high-risk individuals for lifestyle and pharmacological interventions per ESC and ACC/AHA guidelines.^38, 39^ The proportion of SMuRF-less patients varies across racial groups, likely due to genetic factors. In the MINAP registry, whites were more likely to be SMuRF-less, while Asians were less likely (OR: 0.53, 95% CI: 0.50–0.57; *P* < 0.001).^40^

Our study has several strengths, including its position as one of the largest investigations of SMuRF-less STEMI patients in China. By integrating the Chinese CDC database, we were able to follow up on patient outcomes for one year. We included the largest cohort studied to date, along with multiple time-to-surgery variables, allowing for a comprehensive assessment of in-hospital delays in STEMI patients. We identified significant treatment disparities among women between SMuRFs and SMuRF-less, with insufficient preventive awareness emerging as a critical factor contributing to the differences in mortality between groups. These findings underscore the need for more aggressive reperfusion therapy in STEMI patients, particularly women, regardless of the presence of traditional risk factors.

Despite its strengths, this study has limitations. First, as an observational study, while we adjusted for known confounders, unmeasured confounders may exist, and the causal link between SMuRF-less status and mortality remain unclear. Second, SMuRFs classification was based on clinical diagnosis, which may involve omitted or absent prior disease history in some patients. Additionally, some risk factors are continuous, which may create a risk gradient not fully captured.^41^ Third, due to the limited number of variables in the database, some factors affecting the outcomes were not included, which may impact the accuracy of the results. Fourth, as our follow-up focused only on mortality, we lacked data on recurrent myocardial infarction, heart failure, readmissions, and other events, which is a limitation of our analysis. Finally, the inclusion of only certified chest pain centers may exclude hospitals with lower medical levels, introducing potential data bias.

## Conclusion

In conclusion, this multicenter analysis reveals that STEMI patients presenting without conventional modifiable cardiovascular risk factors constitute a substantial proportion (23.1%) of STEMI cases. Notably, this SMuRF-less cohort demonstrated significantly elevated 30-day mortality rates compared to their risk factor-positive counterparts, with male patients exhibiting particularly adverse outcomes during this critical window, the mortality risk profiles converged post-30-day.

## Acknowledgments

The authors thank all hospitals participating in the CCPC for their invaluable contribution to this work.

## Sources of Funding

This work was supported by National Natural Science Foundation of China (82320108001, 82321001, 72274133 and 82270349) and Tianjin Key Medical Discipline (Specialty) Construction Project (Grant No. TJYXZDXK-069C).

## Role of the Funder/Sponsor

The funder had no role in study design, data collection, data analysis, data interpretation, or writing of the report.

## Conflicts of interest

There is no conflict of interest.

## Author contributions

Bin Sun: Conceptualization, Data curation, Formal analysis, Methodology, Writing - original draft. Chunpo Liang: Conceptualization, Supervision, Validation, Writing - original draft. Pengfei Sun: Conceptualization, Supervision, Writing - review & editing. Geru A: Data curation, Methodology, Validation. Liang Zhao: Conceptualization, Methodology. Linjie Li: Formal analysis, Validation, Visualization. Piao Li: Data curation, Methodology. Yongle Li: Supervision, Writing – original draft. Roger Sik-Yin Foo: Supervision, Writing – original draft. Mark Yan-Yee Chan: Supervision, Writing – original draft. Qing Yang: Conceptualization, Supervision, Writing – review & editing. Xin Zhou: Conceptualization, Supervision, Validation, Writing – review & editing.

## Disclosures

The authors declare no competing interests.

## Data availability

The data analysed in this study were based on the China Chest Pain Center database. Individual-level data in the registers can only be accessed through secure servers and only export of aggregated data, as presented in research articles, is allowed as per law. Permission to access data can be made only after fulfilling specific requirements to safeguard the anonymity of the study participants. For these reasons, data cannot be made generally available.

## Supplemental Material

Tables S1, Figure S1-S3

## Notes

### Competing Interest Statement

The authors have declared no competing interest.

